# Alcohol Consumption Patterns and Sociodemographic Correlates Among US Adults with Cardiovascular Disease: A Cross-Sectional Analysis of All of Us and NHANES

**DOI:** 10.64898/2026.06.06.26355052

**Authors:** Qian Yang, Jinyue Yu, Huiling Zhao, Mengxuan Zou, Yangbo Sun

**Affiliations:** MRC Integrative Epidemiology Unit at University of Bristol, Bristol, UK; Population Health Sciences, Bristol Medical School, University of Bristol, Bristol, UK; Nutrition Group, UCL Great Ormond Street Institute of Child Health, University College London, London, UK; Department of Preventive Medicine, The University of Tennessee Health Science Center, Memphis, TN, USA

**Author notes:** Correspondence to Dr. Yangbo Sun. These authors contributed equally (Qian Yang, Jinyue Yu).

**Keywords:** Cardiovascular disease, Public Health, Alcohol consumption, NHANES

## Abstract

This cross-sectional study aimed to examine the prevalence of alcohol use and its sociodemographic correlates among adults with cardiovascular disease (CVD). We analyzed data from two large US cohorts: the All of Us Research Program (2017–2023) and the National Health and Nutrition Examination Survey (NHANES, 1999–2016). Both CVD diagnosis and past-year alcohol consumption were self-reported. Risky drinking was defined as exceeding moderate drinking or binge drinking (All of Us), or moderate/heavy drinking (NHANES). Multivariable logistic regression was used to exam associations with sociodemographic and lifestyle factors. Among 32,788 current drinkers with CVD in the All of Us cohort, 15% exceeded moderate drinking thresholds and 26% reported binge drinking. Older age, female sex, and higher socioeconomic status were inversely associated with risky drinking, while smoking was positively associated. In NHANES, moderate drinking rose from 47.3% to 57.2% and heavy drinking from 6.7% to 7.2%. Moderate/heavy drinking was positively associated with age <65 but inversely with age ≥65. Higher education and income were linked to moderate drinking, while current smoking was strongly associated with heavy drinking. These results highlight the need to integrate holistic screening for alcohol use, tobacco use, and social context into routine cardiovascular care.

## Introduction

Alcohol consumption is a well-established modifiable risk factor for cardiovascular disease (CVD), linked to arrhythmias, hypertension, stroke, and myocardial infarction.^1,2^ Despite ongoing public health messaging, alcohol remains one of the most widely consumed psychoactive substances globally. In the United States, approximately 60–70% of adults report current alcohol use, and over one quarter engage in binge drinking.^3^ These behaviours have important implications for chronic disease management, particularly among individuals with existing cardiometabolic conditions. Historically, a J-shaped association has been reported to suggest that light-to-moderate alcohol consumption may be inversely associated with cardiovascular risk.^4^ However, this relationship has been increasingly questioned, with recent evidence suggesting that even moderate levels of alcohol intake may elevate the risk of certain cardiovascular outcomes.^5^ For example, in a large-scale pooled analysis of 83 prospective studies, Wood et al. showed that risk for stroke and heart failure increased linearly with alcohol consumption, with no protective threshold identified.^2^ These findings have contributed to revised public health guidelines advocating for reduced alcohol use across all population groups.

Research on alcohol and CVD has often focused on primary prevention or general population cohorts. In the general population, alcohol use tends to follow social gradients. Higher income and education levels are often associated with higher likelihood of any alcohol use but lower risk of harmful or dependent drinking; conversely, individuals with lower socioeconomic status (SES) may be less likely to drink overall but more likely to engage in risky patterns when they do.^6,7^ These patterns reflect broader disparities in access to healthcare, stress exposure, and structural disadvantage. Smoking history is another key behavioural factor that may cluster with alcohol use. Smoking and alcohol often co-occur due to shared social and psychological risk factors.^8^ Among individuals with CVD, continued smoking is a major barrier to secondary prevention. A cross-sectional analysis of the All of Us dataset in cancer survivors found that over 75% reported current drinking, with a notable proportion engaging in binge drinking.^9^ However, no comparable analysis has been conducted among adults with CVD, despite their higher baseline risk for alcohol-related complications. Moreover, little is known about how socioeconomic status and smoking interact with alcohol use in this context.

Identification of sociodemographic patterns of alcohol use in this population is essential for developing equitable prevention and counselling strategies. Thus, we used two large and complementary datasets—All of Us and the National Health and Nutrition Examination Survey (NHANES)—to examine patterns of alcohol consumption among adults with self-reported CVD in the United States. Specifically, we aimed to estimate the prevalence of current drinking, exceeding moderate drinking thresholds, and binge drinking; and to assess sociodemographic correlates including age, sex, race/ethnicity, income, education, and smoking status. Moreover, we compare patterns across nationally representative (NHANES) and precision-medicine oriented (All of Us) cohorts.^10^

## Results

### All of Us

Among 45 105 adults with CVD (mean [SD] age, 64.0 [13.4] years; 52.9% women), 32788 (72.7%) reported current alcohol use. Among current drinkers, 15.4% exceeded moderate drinking thresholds, and 26.1% reported binge drinking.

Older age was associated with higher risk of exceed moderate drinking or binge drinking (Table 1). Women were substantially less likely than men to exceed moderate drinking or binge drinking (Table 1). Compared with non-Hispanic White individuals, Hispanic participants had higher odds of both exceeding moderate drinking (OR, 1.29; 95% CI, 1.13–1.43) and binge drinking (OR, 1.23; 95% CI, 1.10–1.37). Non-Hispanic Black individuals had elevated odds of binge drinking (OR, 1.22; 95% CI, 1.11–1.37), while participants identifying as other races were less likely to report either behaviour.

**Table 1.** Odds ratio of current drinking among patients with cardiovascular disease in All of Us.

| Characteristic |  | Exceeding moderate drinking | Binge drinking |
| --- | --- | --- | --- |
|  |  | *Odds ratio (95% confidence interval)* |  |
| Age (years) | <65 | 1 [Reference] | 1 [Reference] |
|  | ≥65 | 0.96 (0.96, 0.96) | 0.95 (0.95, 0.96) |
| Sex | Men | Reference | Reference |
|  | Women | 0.42 (0.39, 0.45) | 0.48 (0.46, 0.51) |
| Education | Below high school | Reference | Reference |
|  | High school diploma or GED | 0.76 (0.62, 0.94) | 0.77 (0.64, 0.94) |
|  | Some college | 0.60 (0.49, 0.73) | 0.60 (0.50, 0.72) |
|  | College | 0.49 (0.40, 0.60) | 0.48 (0.40, 0.58) |
| Smoking status | Never | Reference | Reference |
|  | Former | 2.17 (2.02, 2.34) | 1.62 (1.53, 1.72) |
|  | Current | 3.23 (2.91, 3.57) | 2.27 (2.07, 2.49) |

Abbreviation: GED, General Educational Development certification.

As shown in Table 1, educational attainment showed a strong inverse association. Compared to individuals with less than a high school education, those with a high school diploma or General Educational Development certification had lower odds of exceeding moderate drinking and binge drinking. The odds were lower still among those with some college or a college degree. Compared to individuals with annual household income below $35,000, those earning $75,000–$150,000 had lower odds of exceeding moderate drinking (OR, 0.86; 95% CI, 0.76–0.98). According to Table 1, former smokers had over twice the odds of exceeding moderate drinking (OR, 2.17; 95% CI, 2.02–2.34) and 62% higher odds of binge drinking (OR, 1.62; 95% CI, 1.53–1.72) compared to never smokers. Current smokers had even greater odds: over three times higher for exceeding moderate drinking (OR, 3.23; 95% CI, 2.91–3.57) and approximately 2.3 times higher for binge drinking (OR, 2.27; 95% CI, 2.07– 2.49).

### NHANES

Among adults with cardiovascular disease (CVD) in NHANES 1999–2016, alcohol use patterns showed significant temporal trends (Table 2). The proportion of non-drinkers declined from 46.0% in 1999–2000 to 35.6% in 2015–2016, while moderate drinking increased from 47.3% to 57.2% (peak 62.8% in 2013–2014). Heavy drinking rose slightly from 6.7% to 7.2%.

**Table 2.** Trend in alcohol use among adults with CVD, NHANES 1999-2016.

| Alcohol use | 1999-2000 | 2001-2002 | 2003-2004 | 2005-2006 | 2007-2008 | 2009-2010 | 2011-2012 | 2013-2014 | 2015-2016 |
| --- | --- | --- | --- | --- | --- | --- | --- | --- | --- |
| None | 46.0 | 43.7 | 51.5 | 43.3 | 44.4 | 36.9 | 34.6 | 34.2 | 35.6 |
| Moderate<br>drinking | 47.3 | 50.4 | 43.1 | 50.0 | 49.3 | 57.2 | 55.4 | 62.8 | 57.2 |
| Heavy<br>drinking | 6.7 | 6.0 | 5.5 | 7.1 | 6.3 | 5.9 | 10.0 | 3.1 | 7.2 |

As shown in Table 3, demographic factors were strongly associated with alcohol use. Moderate and heavy drinking increased with age before 65 years but inversely correlated after 65 years. Sex also emerged as a key factor. Males accounted for 63.6% of heavy drinkers, compared to moderate (34.8%) and non-drinking (30.8%) groups. Regarding race/ethnicity, non-Hispanic Blacks constituted the largest proportion across all categories, while non-Hispanic White individuals had the lowest representation in each group. Besides, higher education and poverty-income ratio were positively linked to moderate drinking (p<0.001), with 55.6% of moderate drinkers having high school qualification or above and 31.9% having a poverty-income ratio ≥4. Current smokers dominated heavy drinking (46.3%), while never smokers made up 59.3% of non-drinkers. Adults with ≥1200 MET-min/week of physical activity were more likely to be moderate (39.3%) or heavy drinkers (44.0%) than non-drinkers (25.7%). Higher BMI showed a relatively weak but significant growing proportion across all three drinking categories (p=0.01). Heavy drinkers had the lowest HEI-2010 score compared to moderate and non-drinkers.

**Table 3.** Characteristics associated with alcohol use patterns, NHANES 1999-2016.

| Characteristics | Non<br>drinking | Moderate<br>drinking | Heavy<br>drinking | p |
| --- | --- | --- | --- | --- |
| Age (years), % |  |  |  | <0.001 |
| 20-29 | 1.5 | 2.0 | 3.5 |  |
| 30-39 | 2.3 | 4.6 | 5.1 |  |
| 40-49 | 7.9 | 11.3 | 21.5 |  |
| 50-64 | 22.3 | 31.8 | 36.3 |  |
| ≥65 | 66.0 | 50.2 | 33.6 |  |
| Sex, male (%) | 30.8 | 34.8 | 63.6 | <0.001 |
| Race/ethnicity, % |  |  |  | <0.001 |
| Non-Hispanic White | 8.5 | 6.9 | 6.7 |  |
| Non-Hispanic Black | 71.5 | 79.0 | 75.8 |  |
| Hispanic | 13.8 | 9.4 | 14.1 |  |
| others | 6.2 | 4.7 | 3.5 |  |
| Education, % |  |  |  | <0.001 |
| Less than high school | 33.8 | 19.7 | 24.9 |  |
| High school | 28.6 | 24.8 | 26.3 |  |
| More than high school | 37.6 | 55.6 | 48.9 |  |
| Ratio of family income to<br>poverty, % |  |  |  | <0.001 |
| <1 | 18.7 | 12.5 | 18.1 |  |
| 1-<2 | 32.1 | 20.4 | 24.3 |  |
| 2-<4 | 26.8 | 28.5 | 25.0 |  |
| ≥4 | 14.0 | 31.9 | 28.7 |  |
| missing | 8.5 | 6.7 | 3.9 |  |
| Smoking status, % |  |  |  | <0.001 |
| Non-smoker | 59.3 | 31.6 | 15.9 |  |
| Ever smoker | 26.8 | 44.6 | 37.7 |  |
| Current smoker | 13.9 | 23.7 | 46.3 |  |
| Physical activity level<br>(MET-min/week), % |  |  |  | <0.001 |
| <600 | 64.0 | 48.3 | 45.7 |  |
| 600-1199 | 10.3 | 12.5 | 10.2 |  |
| ≥1200 | 25.7 | 39.3 | 44.0 |  |
| Body mass index, % |  |  |  | 0.01 |
| Normal/underweight | 21.6 | 22.2 | 25.2 |  |
| Overweight | 30.2 | 33.6 | 32.3 |  |
| Obesity | 43.6 | 41.9 | 42.3 |  |
| missing | 4.6 | 2.2 | 0.3 |  |
| HEI-2010 score | 50.4 | 50.5 | 47.0 | 0.01 |

## Discussion

In this nationally diverse cohort, we found that most adults with CVD reported current alcohol use, and a substantial proportion engaged in risky drinking behaviours. Specifically, over 15% of current drinkers exceeded moderate drinking thresholds, and more than 26% reported binge drinking. Our results also showed that in addition to age, sex and ethnicity, alcohol misuse was also associated with socioeconomic disadvantage and smoking history. Individuals with lower income and education levels had higher rates of excessive and binge drinking, highlighting the intersection between alcohol use and social determinants of health. Moreover, both current and former smokers were significantly more likely to engage in risky drinking, suggesting persistent behavioural clustering of CVD-related risk factors.

These findings are broadly consistent with prior research in other clinical populations, including cancer survivors in the All of Us cohort,^13^ where 77.7% reported current drinking and 23.8% binge drinking. Moreover, in a nationally representative sample of myocardial infarction survivors, roughly 20% reported heavy episodic drinking in the past month.^15^ These studies suggest that clinical diagnoses may not meaningfully reduce risky alcohol consumption, high-risk alcohol behaviours are common even among individuals with serious chronic illnesses, particularly when other factors like stress, social norms, or substance use co-occur. In our analysis, the overall prevalence of alcohol use among CVD patients appears slightly lower, but the rate of risky patterns such as binge drinking remains high, suggesting that while some individuals may reduce alcohol use post-diagnosis, others continue or adopt harmful drinking patterns.

Importantly, our results add to a growing body of evidence highlighting socioeconomic gradients in alcohol use patterns. We found that higher income and education were associated with lower odds of exceeding moderate or binge drinking thresholds. This differs somewhat from population-level patterns, where higher SES is typically associated with more drinking, but less harm. In people with CVD, our data suggest that the harms of alcohol may be more concentrated among socially disadvantaged groups. These findings are consistent with the “alcohol harm paradox,” where people of lower SES experience more alcohol-related harm despite drinking the same or less than more advantaged groups.^16^

As for the relationship between smoke and drinking, we found both former and current smokers had substantially elevated odds of exceeding moderate drinking or binge drinking. This reflects longstanding evidence that tobacco and alcohol use are closely linked behaviours, often initiated together and maintained via overlapping neurobiological and social pathways.^17^ In a pooled analysis of U.S. adults, concurrent use of alcohol and tobacco was strongly associated with increased cardiovascular and cancer risk.^18^ Among individuals with existing CVD, this behavioural clustering suggests the need for integrated intervention strategies.

In addition, the trends observed in NHANES from 1999 to 2016 further contextualise our cross-sectional findings. During this period, the prevalence of moderate drinking increased among adults with CVD, while non-drinking declined. Although heavy drinking remained relatively stable, this shift raises concerns that drinking is becoming increasingly normalised, even in high-risk medical populations. Importantly, this shift occurred despite decades of clinical guidance advising alcohol moderation in CVD prevention and management.

Several possible mechanisms may underlie the persistence of risky drinking in this group. These include psychological distress following diagnosis, lack of awareness about alcohol-CVD links, or perceived social norms that minimise the risks of alcohol. Studies had showed high prevalence of depressive symptoms after CVD diagnosis and links distress to adverse health behaviours,^19-21^ and Boden and Fergusson had reported psychological distress following a cardiovascular diagnosis, including depression and anxiety, may contribute to continued alcohol use as a coping strategy.^22^ In qualitative studies, patients with CVD have reported mixed beliefs about alcohol, with some viewing moderate drinking as beneficial for stress relief or social wellbeing.^23^ This highlights the need for patient-centred counselling approaches that engage with personal beliefs and experiences, rather than simply delivering generic advice.

This study has several strengths. The use of two complementary data sources strengthens the robustness and generalisability of our findings. All of Us offers detailed phenotypic and longitudinal data, including diverse racial and ethnic groups often underrepresented in research. NHANES provides a nationally representative sample with longstanding methodological rigour. Although measurement approaches differed slightly, we observed consistent associations between risky drinking and sociodemographic factors in both datasets. More importantly, these datasets provide a more complete picture of alcohol behaviours across different population segments and research contexts. For example, while prevalence estimates differed slightly between cohorts, the sociodemographic patterns were broadly consistent, reinforcing the credibility of observed associations.

Our findings have direct implications for clinical care. Alcohol screening is often underemphasised in CVD management compared to blood pressure, lipids, or glycaemic control. Yet our results suggest that alcohol use is both prevalent and patterned by modifiable social and behavioural risk factors. Routine alcohol screening, using brief tools such as AUDIT-C or single-item questions, could be feasibly incorporated into cardiovascular clinics. Moreover, clinicians should consider broader context, including smoking status, education, and income when assessing alcohol risk. Individuals facing social disadvantage may require tailored counselling approaches that acknowledge economic and psychological constraints.

From a policy standpoint, our findings highlight the need to integrate behavioural health and cardiovascular care. Risky drinking and tobacco use are often treated in silos, yet they co-occur in high-risk populations and are linked to shared social determinants.^24^ Integrated behavioural interventions, delivered through primary care or community health settings, may be more effective than isolated strategies. In addition, public health campaigns targeting alcohol use should avoid generic messaging and instead incorporate culturally and socioeconomically sensitive framing to address health inequities. Future research should also explore longitudinal changes in alcohol behaviour following CVD diagnosis, as well as potential mediators such as stress, medication adherence, or social support. The role of digital health tools and remote counselling may also warrant investigation, particularly for underserved populations with limited access to specialty care.

The study has several limitations. First, alcohol use was self-reported, which may lead to underreporting, particularly among individuals with known health conditions like CVD. Social desirability bias and recall bias could affect accuracy, although such limitations are inherent to most large-scale surveys. Second, our cross-sectional design precludes causal inference. It is unclear whether alcohol use preceded or followed CVD diagnosis. Some individuals may have reduced alcohol consumption after diagnosis, while others may increase use in response to stress or lifestyle disruption. Third, the data did not allow us to distinguish between different types of alcoholic beverages or drinking contexts (e.g., solitary vs. social drinking), which may have differing implications for health. Fourth, although we adjusted for several key covariates, residual confounding from unmeasured variables such as mental health, medication use, or healthcare access remains possible.

## Materials and Methods

### Participants

The All of Us Research Program recruited 453,000 participants through a network of healthcare provider organizations, community centres, and direct volunteer enrolment across the U.S. from 2018 to 2021 ^10,11^. It is a multi-centre study, and longitudinal EHR were linked to participant-provided data. Participation is voluntary, with informed consent obtained from all individuals. All of Us adheres to strict ethical guidelines, overseen by institutional review boards and complies with federal regulations to protect participant privacy and data security. Adults with a self-reported CVD diagnosis who completed the Lifestyle survey were included. Our cross-sectional analysis used data from 6 May 2017 to 1 October 2023. The University of Tennessee Health Science Center institutional review board determined that this analysis was exempt from review and informed consent given the use of deidentified data.

Our study also consisted of participants from the NHANES 1999-2016. NHANES is a nationally representative health survey of the noninstitutionalized US population based on age, gender, and race/ethnicity. Since 1999-2000, NHANES surveys have been organized in 2-year cycles; each cycle consists of approximately 10,000 participants, and collects a wide range of data on demographics, diet, disease history, and health behaviours ^12^. All participants gave written informed consent. The study protocol was approved by the NCHS Research Ethics Review Board. Our cross-sectional analysis included 7792 adults with self-reported CVD diagnosis. After excluding participants with missing information on alcohol use, a final sample of 3627 participants were included in the analysis. The data was accessed for analysis from March 01 2025-Feb 11 2026.

### Drinking behaviours

In All of Us, we followed the previous study conducted among cancer patients to define drinking behaviours ^13^. Current drinking was defined as any alcohol use in the past year. Risky drinking behaviours included (1) exceeding moderate drinking (>2 drinks on a typical day) and (2) binge drinking (≥6 drinks on one occasion).

In NHANES, an alcohol use questionnaire was used to collect information about lifetime and current use (past 12 months) of alcohol among participants. Moderate drinking was defined as up to one drink per day for women and up to two drinks per day for men. while heavy drinking was defined as more than one drink per day for women and more than two drinks per day for men.

### Definitions of demographic, socioeconomic and lifestyle factors

In All of Us, age and sex were self-reported via the “The Basics” module; race/ethnicity was multi-selectable including White, Black/African American/African, Asian, Native American/Alaska Native, Middle Eastern/North African, Native Hawaiian/other Pacific Islander and Hispanic/Latino/Spanish; education was stratified into five levels including below high school, high school graduate/GED, 1–3 years of college/Associate’s degree/technical school, college graduate, and advanced degree; income was categorized by pre-tax household income intervals into six groups including <$25,000; $25,000–<$35,000; $35,000–<$50,000; $50,000–<$75,000; $75,000–<$100,000; ≥$100,000; smoking status (never/former/current) was defined by lifetime cigarette use (≥100 cigarettes) and current frequency via “the Lifestyle” module.

In NHANES, age (AGEDRS) and sex (RIAGENDR) were questionnaire-verified; race/ethnicity was fixed single-select (RIDRETH1/RIDRETH3). Education was coded by DMDEDUC2 (adults) or DMDEDUC3 (adolescents). Income was measured by Poverty-Income Ratio (PIR), which is calculated as the ratio of the total income of a family to the poverty line. Lower PIR indicates poorer socioeconomic status. Smoking status was determined by SMQ020 (lifetime use) and SMQ040 (current frequency). We also considered physical activity, body mass index, and HEI-2010 score. Physical activity was measured via self-reported questionnaires about the number of days per week and the average time spent on each type of physical activity. Body mass index was calculated by weight over height, which were measured using standardized instruments in the NHANES survey. HEI-2010 Score is also called Healthy Eating Index-2010, measured based on data from 24-hour dietary recalls about types and quantities of foods consumed by participants. Higher HEI-2010 Score typically indicates a healthier diet.

### Statistical analysis

In All of Us, we used multivariable logistic regression to assess associations of risky drinking behaviours with the above demographic, socioeconomic and lifestyle factors. For the analysis using NHANES, we followed the NHANES Analytic Guidelines to account for sample weights to obtain variance estimation in the data. Therefore, our results are generalizable to the noninstitutionalized US population. Characteristics of the study sample were weighted and are presented as means for continuous variables or percentages for categorical variables. According to the NHANES analytic tutorials, generalized linear models were used to compare differences in continuous variables, and χ^2^ tests were used in categorical variables.

All analyses were conducted in SAS. This study followed STROBE guidelines,^14^ used deidentified data, and was exempt from IRB review.

## Conclusions

Across two large US population-based cohorts, the majority of adults with established cardiovascular disease reported current alcohol consumption, and a substantial proportion met criteria for risky drinking patterns. Risky drinking was inversely associated with older age, female sex, and higher socioeconomic status, and showed a strong positive correlation with current smoking. These findings underscore the critical need to integrate systematic alcohol screening and intervention into routine cardiovascular care, with particular attention to socially disadvantaged populations and individuals who continue to smoke. Comprehensive lifestyle strategies for chronic disease prevention must address alcohol use, tobacco use, and the broader social determinants of health in an integrated, simultaneous manner.

## Data Availability

The data are available to authorized users with controlled tier access through the All of Us Researcher Workbench at https://www.researchallofus.org/data-tools/workbench/. NHANES data along with documents on the survey methods and other information are publicly available at https://www.cdc.gov/nchs/nhanes/. Further information and requests for resources and reagents should be directed to and will be fulfilled by the lead contact, Yangbo Sun.

https://www.cdc.gov/nchs/nhanes/

## Author contributions

Dr Sun has full access to the data in this study and takes responsibility for the integrity of the data and the accuracy of the data analysis. Concept and design/ Administrative, technical, or material support: Sun, Yang. Acquisition, analysis, or interpretation of data/ Statistical analysis/ Supervision: Sun. Drafting of the manuscript: Yu, Zhao. Critical review of the manuscript for important intellectual content: All authors.

## Funding

This research received no external funding.

## Acknowledgments

The authors would like to thank participants and staff members of the All of Us Research Program and NHANES.

## Competing Interests Statement

The authors declare no competing interests.

## Notes

### Competing Interest Statement

The authors have declared no competing interest.

### Funding Statement

The author(s) received no specific funding for this work.

### Author Declarations

This study is a secondary cross-sectional analysis of publicly available, de-identified population data from the All of Us Research Program and the National Health and Nutrition Examination Survey (NHANES). No direct recruitment of human subjects, no invasive procedures or interventions were performed. Therefore, ethical approval from an institutional review board (IRB) was not required. All original data were ethically approved by their respective oversight bodies and made publicly accessible in compliance with ethical guidelines and data use agreements.

